# Characteristics and outcomes of hospitalized adult COVID-19 patients in Georgia

**DOI:** 10.1101/2020.10.23.20218255

**Authors:** Tengiz Tsertsvadze, Marina Ezugbaia, Marina Endeladze, Levani Ratiani, Neli Javakhishvili, Lika Mumladze, Manana Khotchava, Maiko Janashia, Diana Zviadadze, Levan Gopodze, Alex Gokhelashvili, Revaz Metchurchtlishvili, Akaki Abutidze, Nikoloz Chkhartishvili

## Abstract

**Objective:** Describe presenting characteristics of hospitalized patients and explore factors associated with in-hospital mortality during the first wave of pandemic in Georgia.

**Methods:** This retrospective study included 582 adult patients admitted to 9 dedicated COVID-19 hospitals as of July 30, 2020 (72% of all hospitalizations). Data were abstracted from medical charts. Factors associated with mortality were evaluated in multivariable Poisson regression analysis.

**Results:** Among 582 adults included in this analysis 14.9% were 65+ years old, 49.1% were women, 59.3% had uni- or bi-lateral lung involvement on chest computed tomography, 27.1% had any co-morbidity, 13.2% patients had lymphopenia, 4.1% had neutophilosis, 4.8% had low platelet count, 37.6% had d-dimer levels of >0.5 mcg/l. Overall mortality was 2.1% (12/582). After excluding mild infections, mortality among patients with moderate-to-critical disease was 3.0% (12/399), while among patients with severe-to-critical disease mortality was 12.7% (8/63). Baseline characteristics associated with increased risk of mortality in multivariate regression analysis included: age ≥65 years (RR: 10.38, 95% CI: 1.30-82.75), presence of any chronic co-morbidity (RR: 20.71, 95% CI: 1.58-270.99), lymphopenia (RR: 4.76, 95% CI: 1.52-14.93), neutrophilosis (RR: 7.22, 95% CI: 1.27-41.12), low platelet count (RR: 6.92, 95% CI: 1.18-40.54), elevated d-dimer (RR: 4.45, 95% CI: 1.48-13.35), elevated AST (RR: 6.33, 95% CI: 1.18-33.98).

**Conclusion:** In-hospital mortality during the first wave of pandemic in Georgia was low. We identified several risk factors (older age, co-morbidities and laboratory abnormalities) associated with poor outcome that should provide guidance for planning health sector response as pandemic continues to evolve.

## Introduction

Ongoing pandemic of novel coronavirus disease 2019 (COVID-19) showed differential impact on mortality across the European continent, with cumulative mortality ranging from 1 to over 100 death per 100,000 population (1). Georgia is a small Eastern European country that rapidly responded to the pandemic threat by implementing effective public health and clinical measures achieving one of the lowest mortality rates of 1.25 per 100,000 population as of October 5, 2020 (1). The care pathway model included testing with real-time reverse transcriptase polymerase chain reaction (RT-PCR) of suspected cases (contacts of confirmed cases, persons with symptoms suggestive of COVID-19) with rapid hospitalization of all confirmed cases regardless of disease severity. Sixteen “fever” centres have been established for initial triage and RT-PCR diagnostics of symptomatic persons, while in-patient care was provided in 12 dedicated COVID-19 hospitals. The objective of this study was to describe presenting characteristics of hospitalized patients and explore factors associated with in-hospital mortality during the first wave of pandemic.

## Methods

This retrospective study included unselected consecutive patients with RT-PCR confirmed COVID-19 admitted to 9 designated COVID-19 hospitals across the country between February 26 and July 30, 2020, for whom complete outcome data was known (discharged alive or died in hospital).

As of July 30, 2020, 1160 persons with confirmed COVID-19 have been hospitalized in Georgia, among them outcome was available for 952 patients of whom 935 (98.2%) were discharged alive and 17 (1.8%) died. For present analysis complete outcome was available for 678 hospitalized persons (582 adults and 96 children) of whom 666 (98.2%) were discharged alive and 12 (1.8%) died. All lethal outcomes were registered among adults and therefore analysis was limited to persons aged ≥18 years.

Data were extracted from medical charts, including demographic characteristics, symptoms at presentation, vital signs, laboratory assays, presence of chronic co-morbid conditions, complications during hospital stay, admission to intensive care unit (ICU), need for oxygen support, treatment strategy and outcome of hospitalization. Treatment is provided in accordance with the national protocols, which is the “living guideline” regularly updated with emerging evidence. Confirmed cases were classified into mild, moderate, severe and critical disease.

## Results

Among 582 adults included in this analysis the median age was 46 (IQR: 34-58) years, 14.9% were 65+ years old, 49.1% were women. At the time of hospitalization 13.1% patients had no symptoms, 59.3% had uni- or bi-lateral lung involvement on chest CT and 27.1% patients had any chronic co-morbid condition (cardiovascular disease, diabetes, cancer, chronic obstructive pulmonary disease, chronic kidney disease, chronic liver disease). With regard to baseline laboratory values 13.2% patients had lymphopenia, 4.1% had neutophilosis, 4.8% thrombocytopenia, 37.6% had d-dimer levels of >0.5 mcg/l, 14.8% patients had AST levels >40 IU/l and 14.3% had creatinine levels of >100 mcmol/l. Based on baseline characteristics 183 (31.4%) patients were classified as having mild disease, 336 (57.7%) – moderate disease, 53 (9.1%) – severe disease and 10 (1.7%) –critical disease.

Over the median length of stay of 20 (IQR: 16-24) days 12 adult died with mortality of 2.1%. Baseline characteristics associated with increased risk of mortality in multivariate regression analysis included: age ≥65 years (RR: 10.38, 95% CI: 1.30-82.75), presence of any chronic co-morbidity (RR: 20.71, 95% CI: 1.58-270.99), lymphocyte count <1 × 10^9^/l (RR: 4.76, 95% CI: 1.52-14.93), neutrophil count >8 × 10^9^/l (RR: 7.22, 95% CI: 1.27-41.12), platelet count <150 × 10^9^/l (RR: 6.92, 95% CI: 1.18-40.54), d-dimer >0.5 mcg/ml (RR: 4.45, 95% CI: 1.48-13.35), AST >40 IU/l (RR: 6.33, 95% CI: 1.18-33.98) (Table 1).

**Table 1.** Baseline characteristic of study population and factors associated with mortality (n=582)

|  | Total, n (%) | Died, n (%) | Univariate analysis |  | Multivariate analysis |  |
| --- | --- | --- | --- | --- | --- | --- |
|  |  |  | RR (95% CI) | p value | RR (95% CI) | p value |
| **Age categories** |  |  |  |  |  |  |
| 18-65 | 495 (85.1) | 2 (0.4) | 1 |  | 1 |  |
| 65+ | 87 (14.9) | 10 (11.5) | 28.45 (6.34-127.63) | <0.0001 | 10.38 (1.30-82.75) | 0.03 |
| **Gender** |  |  |  |  |  |  |
| Female | 286 (49.1) | 7 (2.4) | 1 |  | 1 |  |
| Male | 296 (50.9) | 5 (1.7) | 0.69 (0.22-2.15) | 0.52 | 1.82 (0.47-7.08) | 0.38 |
| **Symptoms other than fever** |  |  |  |  |  |  |
| Asymptomatic | 76 (13.1) | 1 (1.3) | 1 |  | 1 |  |
| Symptomatic | 506 (97.8) | 11 (2.2) | 1.65 (0.21-12.62) | 0.63 | 0.17 (0.01-4.23) | 0.28 |
| **Temperature** |  |  |  |  |  |  |
| <37 | 252 (43.3) | 4 (1.6) | 1 |  | 1 |  |
| 37-<38 | 271 (46.6) | 4 (1.5) | 0.93 (0.24-3.68) | 0.92 | 0.27 (0.06-1.16) | 0.08 |
| 38+ | 59 (10.1) | 4 (6.8) | 4.27 (1.10-16.59) | 0.04 | 0.25 (0.06-1.17) | 0.08 |
| **Respiratory rate** |  |  |  |  |  |  |
| <24 | 520 (89.3) | 4 (0.8) | 1 |  | 1 |  |
| 24+ | 62 (10.7) | 8 (12.9) | 16.78 (5.20-54.10) | <0.0001 | 1.90 (0.25-14.58) | 0.53 |
| **Oxygen saturation** |  |  |  |  |  |  |
| ≥94 | 530 (91.1) | 6 (1.1) | 1 |  | 1 |  |
| <94 | 52 (8.9) | 6 (11.5) | 10.19 (3.41-30.47) | <0.0001 | 1.69 (0.37-7.75) | 0.50 |
| **Chest CT** |  |  |  |  |  |  |
| No lung involvement | 237 (40.7) | 1 (0.4) | 1 |  | 1 |  |
| Uni- or Bi-lateral involvement | 345 (59.3) | 11 (3.19) | 7.56 (0.98-58.14) | 0.05 | 1.51 (0.09-24.62) | 0.77 |
| **Comorbidities** |  |  |  |  |  |  |
| No comorbidities | 424 (72.9) | 1 (0.2) | 1 |  | 1 |  |
| Any comorbidity | 158 (27.1) | 11 (7.0) | 29.52 (3.84-226.78) | 0.001 | 21.15 (1.60-280.56) | 0.02 |
| **BMI** |  |  |  |  |  |  |
| <25 | 238 (40.9) | 5 (2.1) | 1 |  | 1 |  |
| 25-30 | 283 (48.6) | 4 (1.4) | 0.67 (0.18-2.48) | 0.55 | 0.19 (0.02-1.48) | 0.11 |
| >30 | 61 (10.5) | 3 (4.9) | 2.34 (0.58-9.53) | 0.23 | 1.50 (0.29-7.88) | 0.63 |
| **Hemoglobin** |  |  |  |  |  |  |
| ≥12 g/dl | 521 (89.5) | 8 (1.5) | 1 |  | 1 |  |
| <12 g/dl | 61 (10.5) | 4 (6.6) | 4.27 (1.32-13.77) | 0.02 | 1.67 (0.54-5.14) | 0.37 |
| **White blood count** |  |  |  |  |  |  |
| ≤11 X 10<sup>9</sup>/l | 564 (96.9) | 9 (1.6) | 1 |  | 1 |  |
| >11 X 10<sup>9</sup>/l | 18 (3.1) | 3 (16.7) | 10.44 (3.09-35.36) | 0.002 | 3.29 (0.31-35.26) | 0.33 |
| **Lymphocyte count** |  |  |  |  |  |  |
| ≥1 X 10<sup>9</sup>/l | 505 (86.8) | 6 (1.2) | 1 |  | 1 |  |
| <1 X 10<sup>9</sup>/l | 77 (13.2) | 6 (7.8) | 6.56 (2.17-19.82) | 0.0009 | 4.74 (1.48-15.10) | 0.008 |
| **Neutrophil count** |  |  |  |  |  |  |
| ≤8 X 10<sup>9</sup>/l | 558 (95.9) | 9 (1.6) | 1 |  | 1 |  |
| >8 X 10<sup>9</sup>/l | 24 (4.1) | 3 (12.5) | 7.75 (2.24-26.81) | 0.001 | 7.27 (1.26-41.89) | 0.03 |
| **Platelet count** |  |  |  |  |  |  |
| ≥150 X 10<sup>9</sup>/l | 553 (95.2) | 10 (1.8) | 1 |  | 1 |  |
| <150 X 10<sup>9</sup>/l | 28 (4.8) | 2 (7.1) | 3.94 (0.91-17.18) | 0.07 | 6.99 (1.20-40.61) | 0.03 |
| **D-dimer** |  |  |  |  |  |  |
| ≤0.5 mcg/ml | 363 (62.4) | 2 (0.6) | 1 |  | 1 |  |
| >0.5 mcg/ml | 219 (37.6) | 10 (4.6) | 8.29 (1.83-37.48) | 0.006 | 4.47 (1.51-13.24) | 0.007 |
| **C reactive protein** |  |  |  |  |  |  |
| ≤5 mg/l | 214 (36.8) | 2 (0.9) | 1 |  | 1 |  |
| >5 mg/l | 368 (63.2) | 10 (2.7) | 2.91 (0.64-13.15) | 0.17 | 1.26 (0.17-9.51) | 0.82 |
| **ALT** |  |  |  |  |  |  |
| ≤40 IU/l | 468 (80.4) | 9 (1.9) | 1 |  | 1 |  |
| >40 IU/l | 114 (19.6) | 3 (2.6) | 1.37 (0.38-4.97) | 0.63 | 4.54 (0.77-26.93) | 0.10 |
| **AST** |  |  |  |  |  |  |
| ≤40 IU/l | 496 (85.2) | 7 (1.4) | 1 |  | 1 |  |
| >40 IU/l | 86 (14.8) | 5 (5.8) | 4.12 (1.34-12.68) | 0.01 | 6.34 (1.18-34.13) | 0.03 |
| **Creatinine** |  |  |  |  |  |  |
| ≤100 mcmol/l | 499 (85.7) | 8 (1.6) | 1 |  | 1 |  |
| >100 mcmol/l | 83 (14.3) | 4 (4.8) | 3.01 (0.93-9.76) | 0.07 | 1.88 (0.56-6.31) | 0.31 |

Mortality rates by severity of disease at presentation were: 0.0% (0/183) for mild disease, 1.2% (4/336) for moderate, 3.8% (2/53) for severe disease and 60% (6/10) for critical disease (Table 2). After excluding mild infections, mortality among patients with moderate-to-critical disease was 3.0% (12/399), while among patients with severe-to-critical disease mortality was 12.7% (8/63).

**Table 2.**
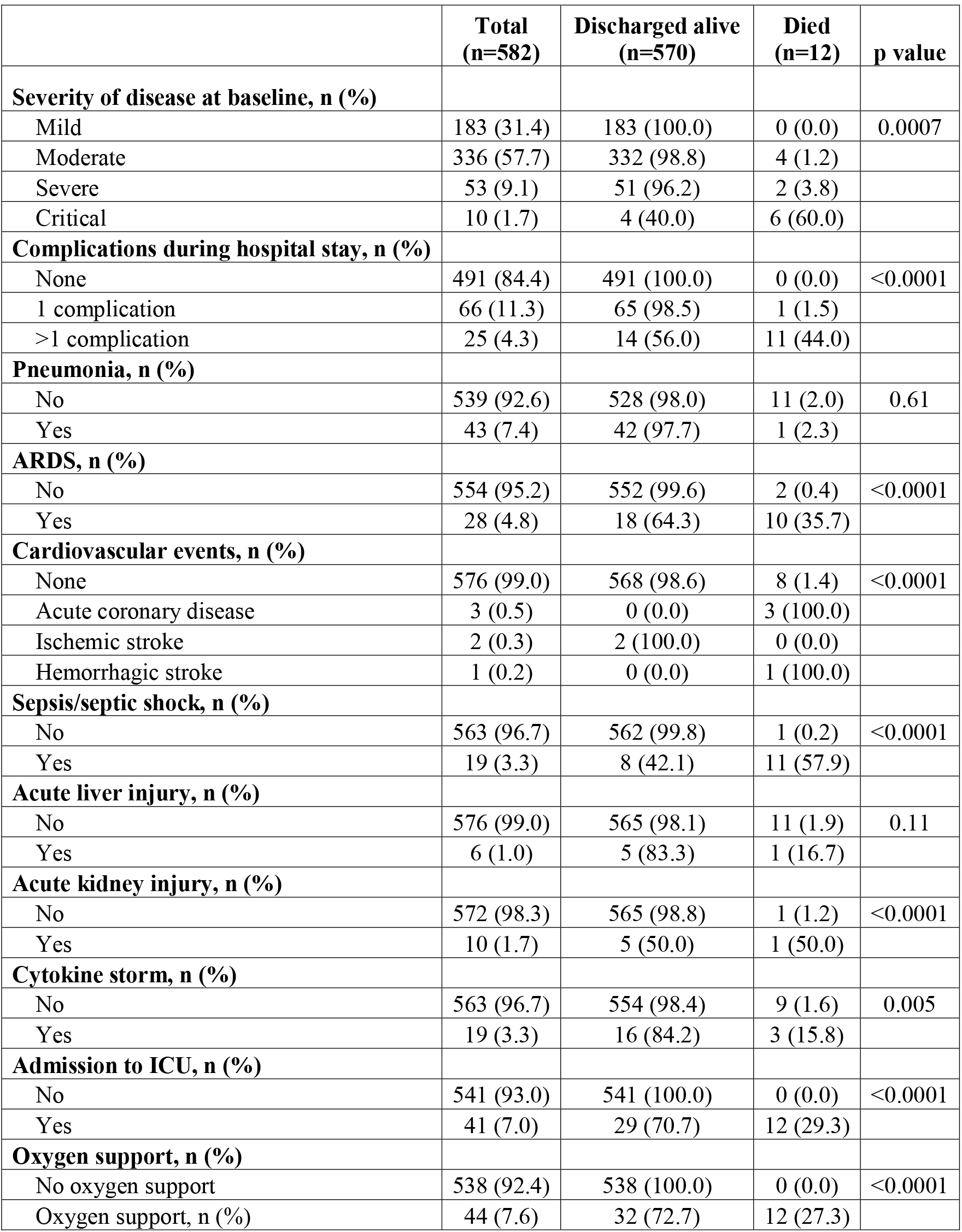
Mortality by severity of disease and complications during hospital stay (n=582)

|  | **Total**<br>**(n=582)** | **Discharged alive**<br>**(n=570)** | **Died**<br>**(n=12)** | **p value** |
| --- | --- | --- | --- | --- |
| **Severity of disease at baseline, n (%)** |  |  |  |  |
| Mild | 183 (31.4) | 183 (100.0) | 0 (0.0) | 0.0007 |
| Moderate | 336 (57.7) | 332 (98.8) | 4 (1.2) |  |
| Severe | 53 (9.1) | 51 (96.2) | 2 (3.8) |  |
| Critical | 10 (1.7) | 4 (40.0) | 6 (60.0) |  |
| **Complications during hospital stay, n (%)** |  |  |  |  |
| None | 491 (84.4) | 491 (100.0) | 0 (0.0) | <0.0001 |
| 1 complication | 66 (11.3) | 65 (98.5) | 1 (1.5) |  |
| >1 complication | 25 (4.3) | 14 (56.0) | 11 (44.0) |  |
| **Pneumonia, n (%)** |  |  |  |  |
| No | 539 (92.6) | 528 (98.0) | 11 (2.0) | 0.61 |
| Yes | 43 (7.4) | 42 (97.7) | 1 (2.3) |  |
| **ARDS, n (%)** |  |  |  |  |
| No | 554 (95.2) | 552 (99.6) | 2 (0.4) | <0.0001 |
| Yes | 28 (4.8) | 18 (64.3) | 10 (35.7) |  |
| **Cardiovascular events, n (%)** |  |  |  |  |
| None | 576 (99.0) | 568 (98.6) | 8 (1.4) | <0.0001 |
| Acute coronary disease | 3 (0.5) | 0 (0.0) | 3 (100.0) |  |
| Ischemic stroke | 2 (0.3) | 2 (100.0) | 0 (0.0) |  |
| Hemorrhagic stroke | 1 (0.2) | 0 (0.0) | 1 (100.0) |  |
| **Sepsis/septic shock, n (%)** |  |  |  |  |
| No | 563 (96.7) | 562 (99.8) | 1 (0.2) | <0.0001 |
| Yes | 19 (3.3) | 8 (42.1) | 11 (57.9) |  |
| **Acute liver injury, n (%)** |  |  |  |  |
| No | 576 (99.0) | 565 (98.1) | 11 (1.9) | 0.11 |
| Yes | 6 (1.0) | 5 (83.3) | 1 (16.7) |  |
| **Acute kidney injury, n (%)** |  |  |  |  |
| No | 572 (98.3) | 565 (98.8) | 1 (1.2) | <0.0001 |
| Yes | 10 (1.7) | 5 (50.0) | 1 (50.0) |  |
| **Cytokine storm, n (%)** |  |  |  |  |
| No | 563 (96.7) | 554 (98.4) | 9 (1.6) | 0.005 |
| Yes | 19 (3.3) | 16 (84.2) | 3 (15.8) |  |
| **Admission to ICU, n (%)** |  |  |  |  |
| No | 541 (93.0) | 541 (100.0) | 0 (0.0) | <0.0001 |
| Yes | 41 (7.0) | 29 (70.7) | 12 (29.3) |  |
| **Oxygen support, n (%)** |  |  |  |  |
| No oxygen support | 538 (92.4) | 538 (100.0) | 0 (0.0) | <0.0001 |
| Oxygen support, n (%) | 44 (7.6) | 32 (72.7) | 12 (27.3) |  |

A total of 91 (15.6%) patients developed complications during hospital stay, including 25 (4.3%) patients developing multiple complications resulting in 11 deaths. Spectrum of complications included 43 (7.4%) new episodes of pneumonia, 19 (3.3%) acute respiratory distress syndrome, 6 (1.0%) cardiovascular events (3 acute coronary disease, 2 ischemic strokes, 1 haemorrhagic stroke), 19 (3.3%) sepsis/septic shock, 6 (1.0%) acute liver injury, 10 (1.7%) acute kidney injury, 19 (3.3%) cytokine storm (Table 2). Forty-one (7.0%) patients required admission to ICU resulting in 12 deaths (29.3% mortality) and 44 (7.6%) patients required oxygen support resulting in 12 deaths (27.3% mortality) (Table 2).

Overall 282 (48.5%) patient received standard of care treatment only, 291 (50.0%) received hydroxycholorquine and 9 (4.8%) - lopinavir/ritonavir. In addition, 338 (58.1%) patients received antibiotics, 52 (8.9%) – corticosteroids, 2 (0.3%) - tocilizumab, 3 (0.5%) – plasmapheresis, 3 (0.5%) blood purification with cytosrobents and 2 (0.3%) transfusion of convalescent plasma.

## Discussion

In this cohort of consecutive hospitalized adult patients with laboratory confirmed COVID-19 we found low in-hospital mortality of 2.1% among total population, 3% - among patients with at least moderate disease and 12.7% among patient with at least severe disease at presentation.

Overall mortality shown in our study is lower than 18.88% reported in recent meta-analysis, even if we limit our analysis to patients with severe/critical disease (2). However, the difference needs to be interpreted with caution in light of varying criteria for hospitalization and rationing of resources in overburdened healthcare systems (3). Reported rates of mortality among patients admitted to ICU or requiring ventilation ranges between 23% and 53% (4-7), with our study showing mortality of 29.3%.

Similar to other studies older age, presence of chronic comorbidities and laboratory abnormalities (leukopenia, neutrophilosis, thromocytopeia and increase in d-dimer) have been significantly associated with mortality (8-11). Interestingly, our data did not show gender-based differences neither in univariate nor multivariate analysis., while reports from other countries universally indicate increased risk of death among men (12).

Predicting outcomes of COVID_19 disease has been a subject of intensive research (13). This is particularly important for Georgia as the country moved from hospitalize all approach to outpatient care of persons with mild disease because of significant increase in diagnosis since September 2020 (1). Defining criteria for hospitalization is crucial for appropriate utilization of healthcare resources without compromising lives. In our study baseline clinical characteristics, such as fever, abnormalities in respiratory rate, oxygen saturation and chest CT imaging showed significant association with mortality only in univariate analysis. As an example, 4 patients in our cohort, initially classified to have moderate disease, had significant deterioration of their condition leading to death. All these 4 patients had pre-existing cardiovascular disease, 3 had elevated d-dimer along with lymphopenia – combination of factors that had been shown to predict progression of disease (14). This suggests that initial clinical presentation may not be sufficient for predicting outcome, it is rather complex issue that requires combinations of several determinants, including laboratory parameters.

Our analysis included 72% of COVID-19 hospitalizations occurring in Georgia during the first wave of pandemic thus ensuring representativeness of national data. Limitations include small sample size and small number of events that affected statistical power. Analysis was limited to laboratory parameters included in basic package of care, while other biomarkers known to be associated with outcome such as albumin, IL-6, ferritin (15), were available only for small subset of patients.

In summary, in-hospital mortality during the first wave of pandemic in Georgia was low. Our study characterized risk factors (older age, co-morbidities and laboratory abnormalities) associated with poor outcome that should provide guidance for planning health sector response as pandemic continues to evolve.

## Data Availability

Data is available upon request that should be made to Dr. Tengiz Tsertsvadze at

